# Evaluation of filtration efficacy of various types of facemasks using ambient and PAO aerosols following with different sterilization methods

**DOI:** 10.1101/2020.10.23.20218073

**Authors:** Amit Kumar, Basundhara Bhattacharjee, D.N Sangeetha, V Subramanian, B Venkatraman

**Affiliations:** Radiological and Environmental Safety Division, Indira Gandhi Centre for Atomic Research, Kalpakkam-603102, India; Biomedical Engineering, SSN collage of Engineering, Kalavakkam-603110, India

**Keywords:** pandemic, N-95, surgical mask, cloth mask, autoclave, dry heat sterilization, H_2_O_2_ sterilization

## Abstract

Due to the ongoing pandemic, various types of facemasks such as N-95, surgical mask and cloth masks are being used as an essential protective measure. The filtration efficiency of these masks were tested before and after sterilization by different methods for two flow rates conditions corresponding to normal breathe rate (20 lpm) and during sneezing (90 lpm). Sterilization techniques used here are autoclaving (30 and 60 minutes), dry oven heating (30 and 60 minutes), gamma irradiation (15 and 25 kGy), hot water washing with and without detergent and immersing in 10% concentration of liquid hydrogen peroxide for 30 minutes. As expected, the N-95 is the greatest filtering efficiency among all the other type face masks. The best method to sterilize N-95 masks without affecting its performance is by using dry heat with temperature ranging from 70-80°C. The cloth masks and surgical mask are performed more or less same for both flow conditions. As an affordable sterilization method hot water washing is highly recommended which does not deteriorate the efficiency of the masks and can be used for the general public. The use of double or triple layer cotton cloth masks in the general public serves fit for the purpose than surgical masks. The surgical mask can be sterilized only few times with the help of dry heat, hot water wash and/or autoclave.

## 1.0 Introduction

Since the outbreak of the novel corona virus disease i.e., COVID-19, one of the essential Personal Protective Equipment (PPE) used in this pandemic is face masks which offers protection against covid-19 through droplet and aerosols transmission. The uses of facemasks have become compulsory in India and other countries across the world. Hence, it is become mandatory to wear a facemask by public and workers in the front line working against Covid19 pandemic. Good quality masks can prevent transmission of virus particles from one person to another. Starting from just 1 case in 30^th^ January 2020, we now have more than 7 million cases as of 15^th^ October 2020 (https://www.covid19india.org/). Frontline workers in India and around the world are facing shortage of masks because of the increase in number of the usage. To solve this crisis, masks are being reused multiple times. But using a used facemask for multiple times without killing the pathogens (if any) can result in allowing any virus or bacteria on its surface to directly or indirectly enter into our respiratory system. Here the various sterilization/decontamination processes comes into play. Sterilization can be defined as the procedure of destroying all microorganisms in or on a given environment in order to prevent the spread of infection. Under these scenario, multiple potential methods for sterilization have begun to be explored (Juan et al., 2020).The thermal heat, radiation or chemical agents usually do this. Many potential methods of sterilization have been explored already. They are based on chemical and physical methods. Chemical methods include hydrogen peroxide, chlorine dioxide, bleach, alcohol, soap solution and ethylene oxide, ozone decontamination etc and physical methods include dry/steam heat treatment, UV light sterilization, electron beam and gamma irradiation etc (Kumar et al., 2015; Liao et al., 2020). All above sterilization methods having advantages and disadvantages from one than others (Cramer et. al., 2020; Juan et al., 2020). Some of the methods such as soaking/dipping in alcohol and others damage the fibers of face masks and significantly degrade the filter efficiency. This happens because many masks use fibers that have a static charge which are removed by dipping in alcohol. In the present work, we used some of the methods mentioned above to sterilize the masks and evaluated for filtration efficiency. Those methods include autoclaving for 30 and 60 minutes, gamma irradiation (15 and 25 kGy), dry heat (30 and 60 minutes), detergent wash with hot water, hot water wash and soaking in 10% concentration of hydrogen peroxide for 30 minutes.

In this situation, mostly three types of masks are being used widely in most of the countries namely N-95 (health care professionals), non-woven fabric surgical masks (front line workers) and cloth masks (common publics). While N-95 and surgical masks are being used by the frontline workers during this pandemic, but not all can afford the expensive N-95 face masks, and the public often uses mostly cotton masks as Government of India made it mandatory to wear masks in public places. So the usage of cloth masks by the public has increased rapidly. Cloth masks are preferred for community use because they also reduce the non-biodegradable waste from more use of surgical masks. At this juncture, the reusability of these cloth masks is being carried out, by adopting simple method of sterilization, which is less cost and affordable by the common public. Hence it is very important to study the effectiveness of sterilization by simple methods for these cotton masks. Besides, the effectiveness of different types of cloth masks in blocking respiratory droplets depends on the weave and number of layers. Earlier studies have also shown that the mask efficiency was directly related to the closeness of the mesh and the number of thicknesses of gauge. We have taken two variants of cloth masks, one being two layer cloth mask of same fabric (cotton) and other two layer cloth mask of different fabric (cotton and gada). Since, N-95 is expensive cannot be used by everyone (due to different economic strata of India), a common person can afford cloth mask and surgical mask for protection. The filter efficiencies of these masks were first tested and then were sterilized using the above-mentioned methods and then efficiency was evaluated. The detailed experimental method, results and discussion, conclusion and recommendations are described in this article.

## 2.0 Materials and Methods

The evaluation of Filtration Efficiency (FE) of different types of masks are carried out in a test facility called HEPA Filter Testing Laboratory, Radiological and Environment Safety Division, Indira Gandhi Centre for Atomic Research (IGCAR) consisting of stainless steel cylindrical duct which has provision for fixing the facemask without any air leakage. The upstream and downstream aerosol concentrations are counted and the FE of mask is determined before and after various sterilization methods. The details of the experimental setup, types and quantity of masks, necessary data acquisition systems, sterilization methods and evaluation of FE of masks for ambient and laboratory generated aerosols are explained in this section.

### 2.1 Experimental setup

The schematic diagrame of experimental setup for face mask testing is shown in Fig.1. It consist of a cylindrical tube of 8.5 cm internal diameter (D) with upstream and downstream segment and a middle region to connect facemasks media. The upstream length is more than 15D and the concentration measurements in the upstream and downstream are taken at 5D away from the connection flange to get uniform concentration along the entire duct. The experimental setup also includes aerosols generator, aerosols diagnostic instruments, velocity meter, differential pressure monitor and relative humidity and temperature monitoring system of air stream. The test piece (without cutting the facemask) is connected in between two flanges fitted with O-rings and mounted in the duct and it is perfectly found to be fitted without any air leakage (the mask edges were completely sealed).The desired airflow rates are achieved through suction by using an air displacement pump (Make: M/s. Rotovac and CG commercial motor) and flow controller (M/s. Weiser). The aerosols are sampled before (upstream) and after (downstream) of the specimen when aerosol-air suspension passes through the facemask.

**Fig. 1.**
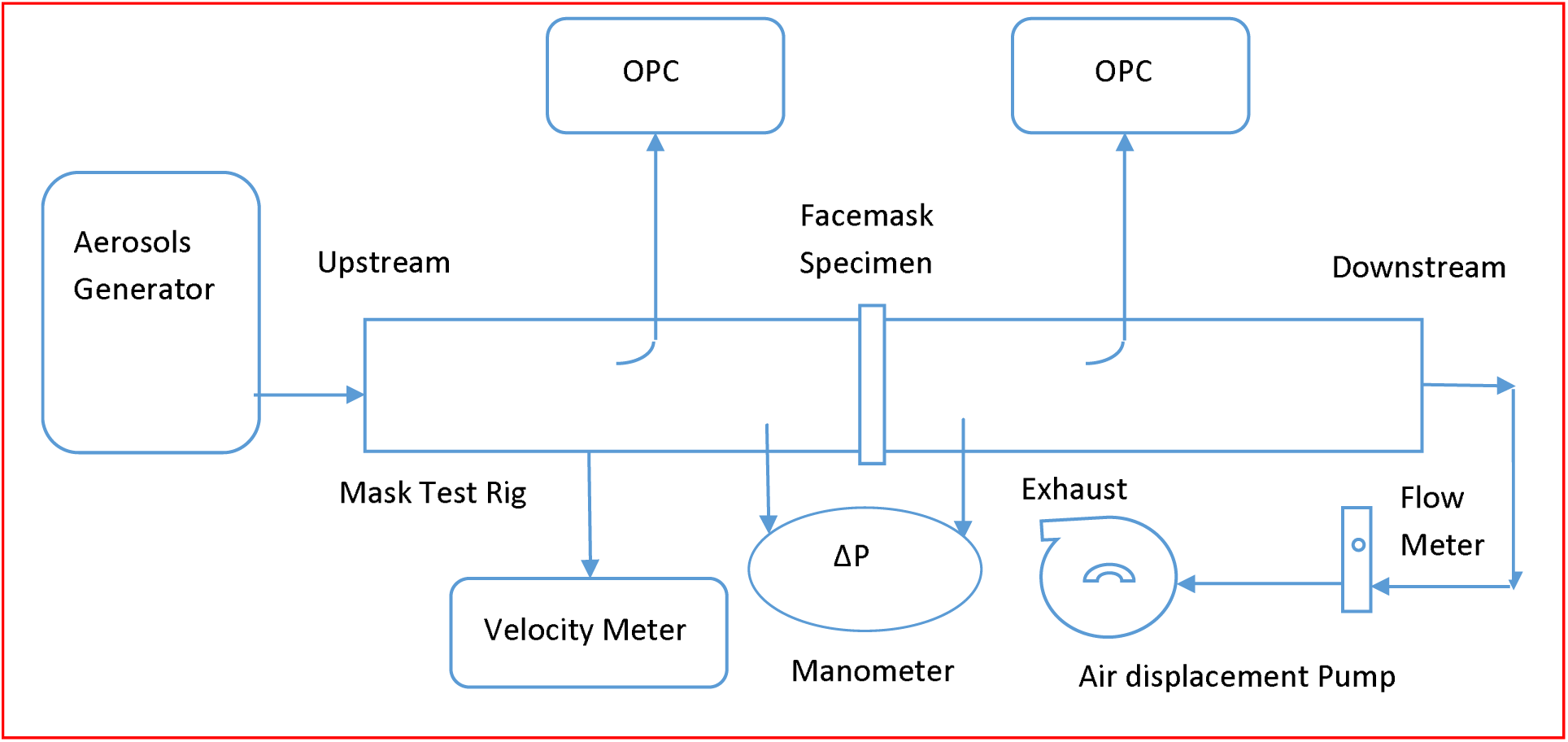
Schematic presentation of experimental setup.

### 2.2 Test aerosols generation and aerosols diagnostic instruments

All types of facemasks are tested for FE with ambient and laboratory generated aerosols. The aerosols suspended in ambient air are continuously introduced at the inlet of the test rig. Poly Alfa Olefin (PAO) aerosols are generated using Laskin-Nozzle aerosol generator (Model and Make: TDA-4B and M/s Air Techniques Internationals, USA). and used as polydispersed test aerosols to test the facemasks. The aerosol sampling is carried out from the upstream and downstream simultaneously using two number of Optical Particle Counter (OPC). The OPC (Model and Make: 1.108 and M/s Grimm Aerosol Technik, Germany)works on the principle of light scattering and measures the particles in the range of 0.3□20 µm. The details of the both instrument working principle and specification can be found elsewhere (Kumar *et al*., 2020 and Usha *et al*., 2019).The reproducibility of the OPC is ± 3% for the entire measuring range of OPC (0.3 to 20 µm). The acceptable accuracy in measurement of aerosols number concentration from OPC is ± 3%.

### 2.3 Types of protective facemasks

The three types of facemasks viz. N-95 (certified facemasks), non-woven fabric (equivalent to surgical mask) and self-made double layer cotton cloths (general public choice during the pandemic) have been tested for particulate filtering efficiency. The types of face masks and their description along with image are summarized in Table1.Five types of certified N-95 facemask commonly used healthcare professionals were selected to examine whether they maintain their performance and integrity after single sterilization. The surgical masks are made of 3 layered gauze having high count gauze (40 x 24) at the outer side and two layers of normal gauze (28 x 24 counts) on the inner side. Two types of double layer cotton masks (same fabric and different fabric) were also investigated. The cotton masks having same fabric are stitched from a bundle cloth made of 2×2 full voile twill weave (two warp threads crossing every two weft thread) having 80 x 80 counts per square inch. The other cloth mask consists of one layer of 2×2 full voile twill weave at the front side and single ply cotton fabric (approx 180GSM) at the rear side. All the facemasks when fitted to the sampler holder care have been taken that masks should not stretch and there should be no leakage from the edges.

### 2.4 Sterilization methods

The following sterilization methods namely, gamma irradiation, dry heat, autoclave (steam), hot water with and without detergent wash and soaking in H_2_O_2_ liquid were carried out for various types of facemasks employed in this study and they are summarized in Table 2. The gamma sterilization of various type of masks is achieved by using gamma irradiation chamber (Model and Make: GC500 and M/s BRIT, India) where the masks are irradiated using Co-60 source (gamma energy 1.17 and 1.33 MeV) for the desired dose level (15 and 25 kGy).The details of gamma irradiation chamber can be found elsewhere (Kumar et al., 2020).

**Table 1.**
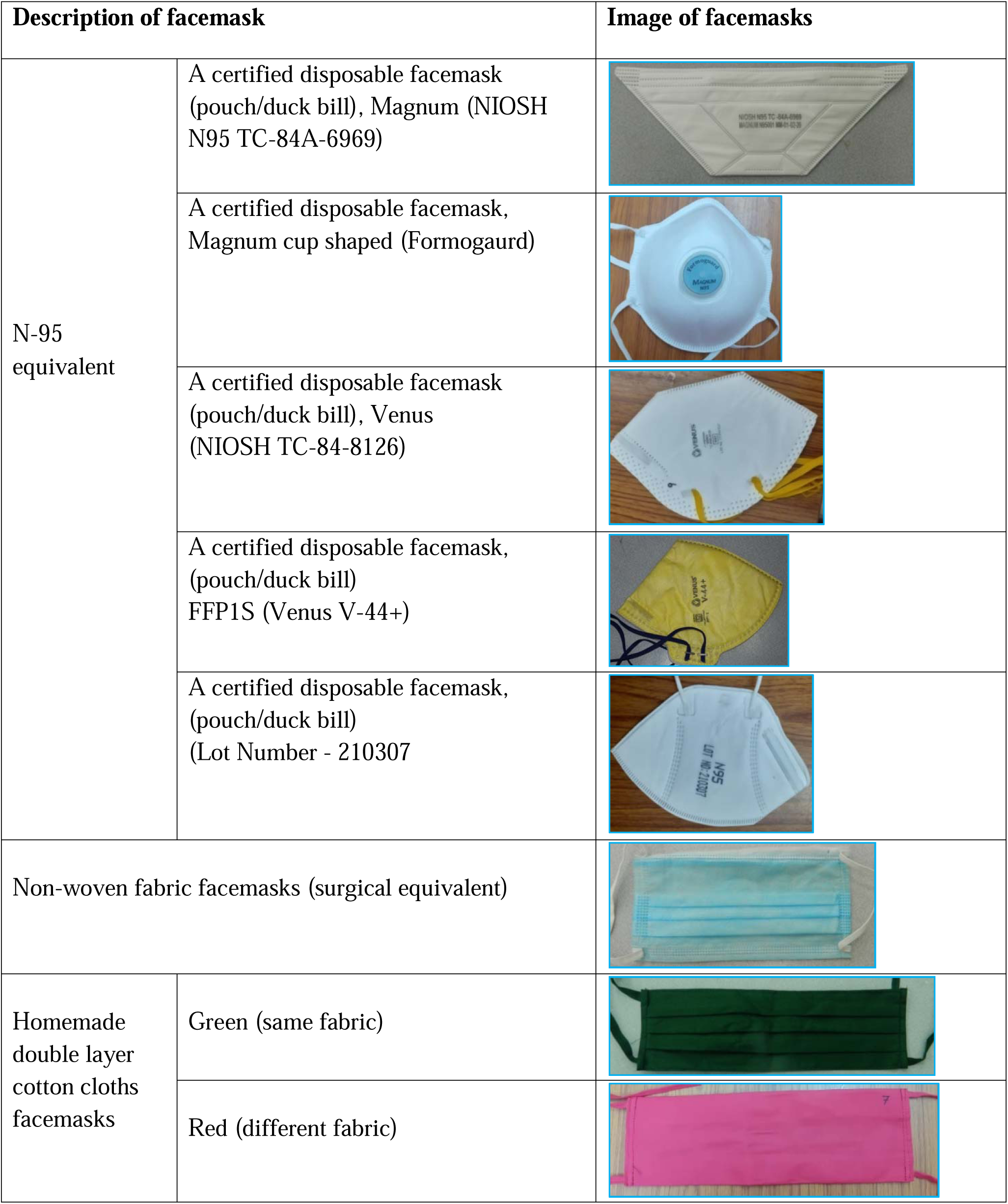
Description and types of facemasks along with their image.

| Description of facemask |  | Image of facemasks |
| --- | --- | --- |
| N-95 equivalent | A certified disposable facemask (pouch/duck bill), Magnum (NIOSH N95 TC-84A-6969) |  |
|  | A certified disposable facemask, Magnum cup shaped (Formogaurd) |  |
|  | A certified disposable facemask (pouch/duck bill), Venus (NIOSH TC-84-8126) |  |
|  | A certified disposable facemask, (pouch/duck bill) FFP1S (Venus V-44+) |  |
|  | A certified disposable facemask, (pouch/duck bill) (Lot Number - 210307) |  |
| Non-woven fabric facemasks (surgical equivalent) |  |  |
| Homemade double layer cotton cloths facemasks | Green (same fabric) |  |
|  | Red (different fabric) |  |

**Table 2.** Different methods used for sterilization of various types of facemasks.

| Types of facemask | Gamma irradiation |  | Autoclave |  | Dry heat |  | Immerse in 10% H<sub>2</sub>O<sub>2</sub> for 30 min | Hot water wash |  |
| --- | --- | --- | --- | --- | --- | --- | --- | --- | --- |
|  | 15 (kGy) | 25 (kGy) | 30 (min) | 60 (min) | 30 (min) | 60 (min) |  | With Detergent | Without Detergent |
| **Magnum (NIOSH)** | ✓ | ✓ | x | x | ✓ | ✓ | ✓ | x | x |
| **Magnum cup** | x | x | ✓ | x | x | x | x | x | x |
| **Venus (NIOSH)** | x | x | ✓ | ✓ | x | x | ✓ | x | x |
| **Venus (V-44+)** | x | x | ✓ | x | x | x | x | x | x |
| **N-95 (210307)** | x | x | ✓ | x | x | x | x | x | x |
| **Surgical** | ✓ | ✓ | ✓ | ✓ | x | x | ✓ | ✓ | ✓ |
| **Cloth (Green)** | ✓ | ✓ | ✓ | ✓ | x | x | x | ✓ | ✓ |
| **Cloth (red)** | x | x | ✓ | ✓ | x | x | x | ✓ | ✓ |

The dry heat decontamination of face masks for 30 and 60 minutes is achieved by hot air oven (Labtherm Scientific Products, Sr. No.0013) with air temperature ranging from 70-80°C. Steam sterilization has been achieved by autoclave machine and sterilization process takes place at a pressure of 103 kPa and temperature of 121°C. Masks were subjected to the autoclave for 30 minutes and 60 minutes of each type of masks (Make: Nat Steel semi-automatic autoclave, Mumbai India). The facemasks sterilization has been also carried out with detergent cum hot water, hot water and 10% concentration of H_2_O_2_ (diluted with distilled water) by soaking for 30 minutes and dried before testing the face masks.

### 2.5 Measurements of differential pressure across the facemask, face velocity, relative humidity and temperature of air stream

The pressure drop or differential pressure (ΔP) was measured using a manometer (Make and model: TSI, USA and 9565P143003) across the tested masks. The measuring range and error in reading in ΔP is −381 to +381 and ±1% of reading + 0.13 mm of H_2_O respectively. The differential pressure is an indicator for condition of comfort and breathability of facemask. The face velocity in the test section is measured with a velocity meter (Make and model: Velocicalc, TSI, USA and 9565P143003). The measuring range and error in reading in air face velocity is 0 □ 50 m s^-1^and ±3% respectively. The K type thermocouple and humidity monitor are used for the measurement of temperature (T) and relative humidity (RH) of air stream respectively. The measuring range along with error in the measurement of T and RH are −10 to 60 °C± 0.3 °C and 5 to 95% ± 3% respectively.

### 2.6 Data acquirement and analysis

The *FE* of three different types of facemask is calculated using the following formula:

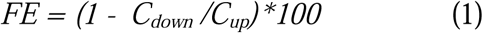

Where, *C*_*up*_ and *C*_*down*_ are the average aerosols number concentration of upstream and downstream respectively for each size bin. The sampled data of each mask has been recorded for about 2 □ 3 min in interval of 6 s from OPC (total sample record 20 □ 30). The *FE* is computed by taking the average of the collected data of upstream and downstream for the tested period and for each filter. The *FE* of the three types of mask is calculated for polydispersed PAO and ambient and aerosols.. The aerosols concentration recorded by instruments are analyzed for each masks (N95, non-woven fabric (surgical mask) and self-made double layer cotton cloths) for five interval (0.3-0.5, 0.5-1.0, 1.0-3.0, 3.0-5.0 and 5.0-10.0 µm) and *FE* were averaged for all similar type masks and tested conditions (like flow rate and sterilization methods). The average *FE* was subjected to one iteration of the Grubbs test with a 95% confidence interval to remove most one outlier. This improves the statistical variation of the data for the calculation of *FE*. Furthermore, the uncertainty associated in *FE* of facemasks has been calculated. Type A uncertainty has been arrived at for repeated measurement and number of face masks tested and Type B uncertainty has been calculated based on the following individual uncertainty (accuracy in measurement, resolution and calibration of instrument). After that, the combined uncertainty has been estimated and then expanded uncertainty derived from combined uncertainty by multiplying the coverage factor (95% confidence level).

## 3.0 Results and discussion

### 3.1 Qualification of experimental setup

Before start the testing of facemasks, the experimental setup has been evaluated. The facemask experimental test rig is tested for zero efficiency condition using ambient aerosols. The upstream and downstream counts are recorded without facemask. The average upstream and downstream aerosol counts as a function of aerosols size is given in Fig. 2. It is observed from the Fig. 2 that, the difference in the upstream and downstream counts ranges 10 - 30 #/L up to aerosol size 0.7 µm and after that difference is very less (1 - 3 #/L) observed.

**Fig. 2.**
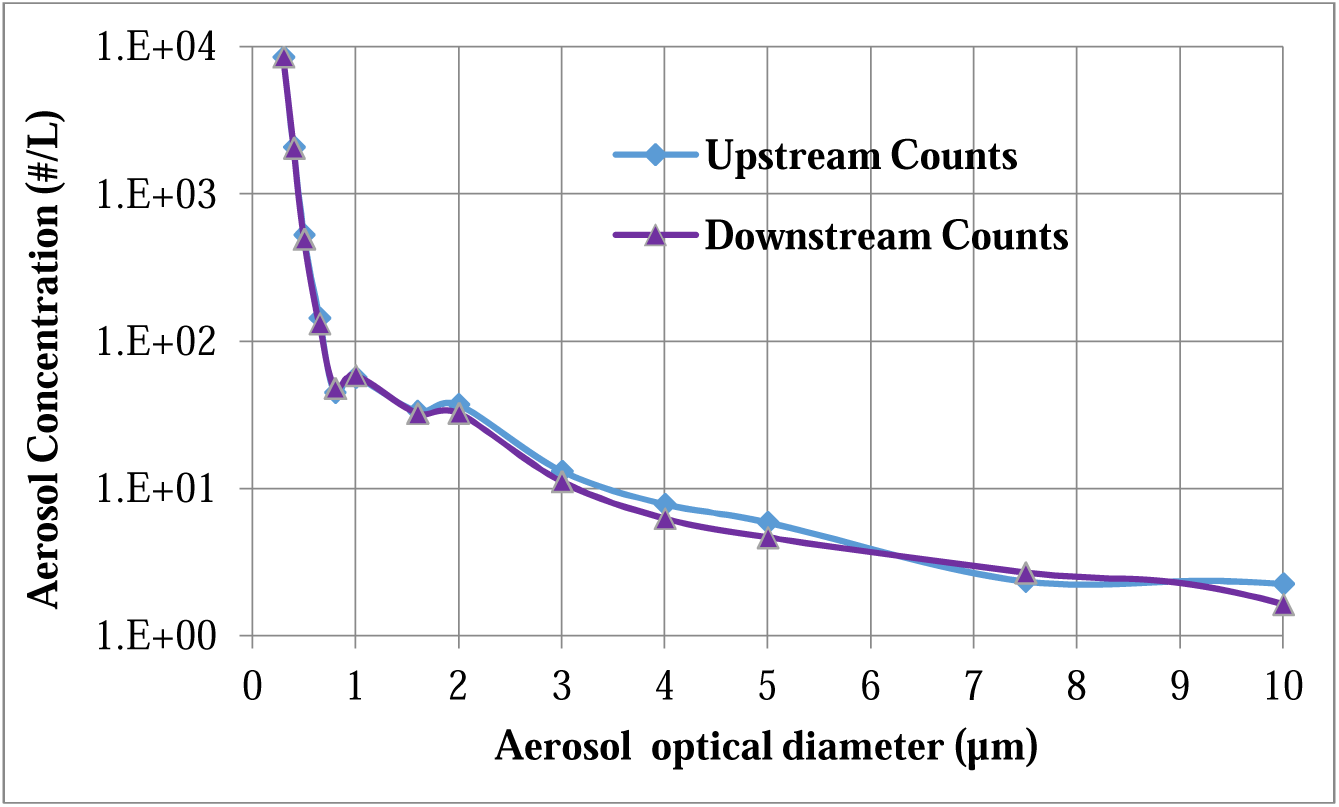
Upstream and downstream average aerosols counts as a function of optical diameter measured by OPC.

The total upstream and downstream aerosols counts are measured by OPC of the test rig without face mask for 20 and 90 lpm flow rate and given in table 3. The zero efficiency (without face mask) with uncertainty of the test rig was tested using ambient aerosols and found to be 2.07±0.55% and 2.35±0.65% for 20 and 90 lpm respectively. The correction factor of zero efficiency has been included in estimation of uncertainty in FE of tested facemask. Further, the measured average aerosol concentration at the upstream (*C*_*up*_) and two-flow rates 20 and 90 lpm for ambient and PAO aerosols are presented in Fig. 3 (A) and (B) respectively. It is observed from Fig. 3 (A) and (B) that, a significantly lower aerosol concentration in the upper size range for both flow conditions, that is, for aerosol greater than 10µm. The data were excluded above this threshold (10 µm) for all results presented in the paper due to the extremely low concentration and this may upsurge statistical error.

**Table 3.** The zero efficiency test of the test rig for ambient aerosols.

| Total average Upstream counts (# L<sup>-1</sup>) | Total average Downstream Counts (# L<sup>-1</sup>) | Efficiency (%) | Average Efficiency + Uncertainty |
| --- | --- | --- | --- |
| **For 20 lpm flow rate** |  |  |  |
| 15,850 | 15312 | 3.39 | 2.07 ± 0.55 |
| 15,298 | 15016 | 1.84 |  |
| 14,932 | 14560 | 2.49 |  |
| 14,016 | 13859 | 1.12 |  |
| 14,631 | 14316 | 2.15 |  |
| **Average : 14,945** | **Average : 14,633** | **Average : 2.07** |  |
| **For 90 lpm flow rate** |  |  |  |
| 15,480 | 15125 | 2.29 | 2.35 ± 0.65 |
| 14,600 | 14421 | 1.23 |  |
| 14,030 | 13724 | 2.18 |  |
| 13,735 | 13298 | 3.18 |  |
| 13,936 | 13539 | 2.85 |  |
| **Average : 14,356** | **Average : 14,009** | **Average : 2.35** |  |

**Fig. 3.**
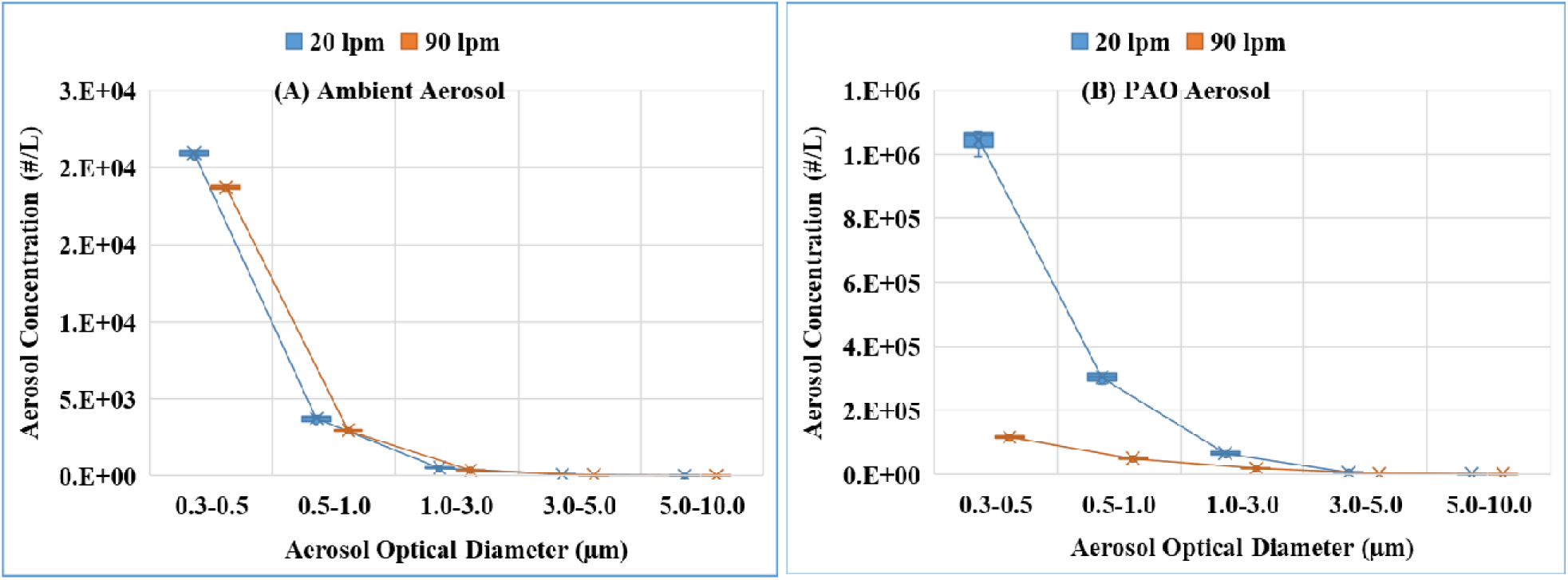
Aerosols concentration as a function of optical diameter of aerosols at a two flow rates, ambient (A) and PAO (B) aerosols.

The three types of facemask viz., N-95, non-woven fabric mask and cloth masks were tested for particulate FE under two conditions 20 and 90 lpm which corresponds to the normal breathing rate and strenuous work load or sneezing/coughing respectively (Adams, 1993 and Lee *et al*., 2019).The surgical mask and cloth mask are mostly rectangular in shape (length is greater than breadth) and N-95 mask in cup/bell shape (it was neither rectangular nor circular).Hence, the entire area of the duct is covered by the facemask and ensured that most of the exposure portion of the mask has been fitted in the test rig and tested. The effective area of the various masks during the test was 56.74 cm^2^.The face area is used for testing of facemask/filter are in line with other research work viz. 78.5 cm^2^ (Heim et al., 2005),17.34 cm^2^ (Kim et al., 2007) and 45 −100 cm^2^ (Konda et al., 2020).The measured ambient air temperature is 27±0.3°C and RH% is 60-70±3%.The challenge aerosols were blown through the mask with a face velocity of 0.058±0.002 and 0.264±0.009 m/s corresponding to 20 ± 0.2 and 90 ± 1.0 lpm flow rates respectively. Before starting the experiment, the tested facemask has been conditioned for about 30 minutes in the same environment.

### 3.2 Differential pressure drop across the facemasks

The measured pressure drop (ΔP) across the various facemasks before and after sterilization is given in Table 4. As expected, the ΔP across all type of facemasks increases with increase of face velocity. The average pressure drop of certified masks viz. Magnum (NIOSH), Magnum Cup shape, Venus (NIOSH), Venus (V-44+) and N-95 (210307) is 3.2 ±0.22, 4.27±0.24, 1.02±0.14, 1.54±0.18 and 1.07±0.15 mm of H_2_O for 20 lpm and ΔP is 26.4±0.47, 24.46±0.43, 7.37±0.21, 5.33±0.21 and 8.10±0.25mm of H_2_O for 90 lpm correspondingly. The Magnum mask has more pressure drop compared to Venus and N-95 (210307) for both flow conditions.

**Table 4.** Measured pressure drop across the facemasks before and after sterilization.

| Type of facemasks | $\Delta P$ (mm of H<sub>2</sub>O) | | | | |
| --- | --- | --- | --- | --- | --- |
|  | Before sterilization |  | After sterilization |  |  |
|  | 20 lpm | 90 lpm | Sterilization | 20 lpm | 90 lpm |
| Magnum (NIOSH) N-95 | 3.2±0.22 | 26.4±0.47 | Dry heat | 3.12±0.23 | 27.02±0.38 |
|  |  |  | □ irradiation | 3.15±0.27 | 26.28±0.37 |
|  |  |  | H<sub>2</sub>O<sub>2</sub> | 3.10±0.25 | 24.28±0.36 |
| Magnum cup N-95 | 4.27±0.24 | 24.46±0.43 | Autoclave | 4.42±0.24 | 25.46±0.44 |
| Venus (NIOSH) N-95 | 1.02±0.14 | 7.37±0.21 | Autoclave | 1.21±0.15 | 8.97±0.23 |
|  |  |  | H<sub>2</sub>O<sub>2</sub> wash | 1.78±0.14 | 12.37±0.26 |
| Venus (V44+) N-95 | 1.54±0.18 | 5.33±0.21 | Autoclave | 1.14±0.14 | 4.36±0.19 |
| N-95 (210307) | 1.07±0.15 | 8.10±0.25 | Autoclave | 1.15±0.16 | 7.16±0.24 |
| Surgical equivalent | 0.26±0.14 | 0.79±0.21 | Autoclave | 0.18±0.13 | 0.45±0.18 |
|  |  |  | □ irradiation | 0.31±0.19 | 0.76±0.22 |
|  |  |  | H<sub>2</sub>O<sub>2</sub> wash | 0.17±0.13 | 0.55±0.19 |
|  |  |  | Hot water | 0.21±0.14 | 0.49±0.17 |
| Green cloth (same fabric) | 0.28±0.15 | 3.41±0.25 | Autoclave | 0.18±0.13 | 2.77±0.14 |
|  |  |  | □ irradiation | 1.31±0.18 | 3.45±0.25 |
|  |  |  | H<sub>2</sub>O<sub>2</sub> wash | 0.15±0.13 | 2.69±0.14 |
| Red cloth (different fabric) | 0.18±0.13 | 2.63±0.19 | Autoclave | 0.23±0.14 | 3.18±0.16 |
|  |  |  | Hot water | 0.23±0.13 | 2.63±0.15 |

The ΔP across Magnum (NIOSH) is remain almost same after dry heat, gamma and H_2_O_2_ sterilization while for Magnum cup shaped mask, the ΔP slightly increased after autoclave sterilization. In the case of Venus (NIOSH) masks, the ΔP is slightly increased, while for V-44+ masks ΔP decreased for both flow conditions. The ΔP across N-95 (210307) masks is slightly increased for low flow rate while slightly increased for high flow rate when compared to the control masks. The average pressure drop of control surgical masks is found to be 0.26±0.14and 0.79±0.21mm of H_2_O for 20 and 90 lpm flow rate. The average ΔP of surgical mask after gamma sterilization is remain same while for other decontamination it is slightly reduced for both flow conditions. Similarly, the average ΔP of controlled green cloth masks (same fabric) is found to be 0.28±0.15 and 3.41±0.25mm of H_2_O for 20 and 90 lpm flow rate respectively. The average ΔP of controlled red cloth masks (different fabric) is found to be 0.18±0.13and 2.63±0.19 mm of H_2_O for 20 and 90 lpm flow rate respectively. The average pressure drop of surgical masks is more than cloth masks and less than certified N-95 masks for both flow rates. It is to be mentioned that the pressure drop across all the two types of cloth decontaminated facemask showed that there is no measurable variations observed i.e. change appears within the error bar values, i.e. there is no physical change in bulk density of face mask threads after decontamination. The pressure drop indicates the condition for usage during breathing and is found to be in the accepted range, 35 and 25 mm of H_2_O for inhalation and exhalation resistance limit respectively (Lin *et al*., 2020).

### 3.3 Evaluation of filtering efficiency of control masks for ambient aerosols of size 0.3 – 10 µm

The FE of various types of control facemasks are determined with ambient aerosols for particle size 0.3 - 10.0 µm, with five size intervals (0.3-0.5, 0.5-1.0, 1.0-3.0, 3.0-5.0 and 5.0-10.0 µm), and for two flow rates and given in Fig.4. It is observed from the Fig.4 that, the mean FE of control surgical masks is 24.72±2.54, 47.97±6.65, 69.51±9.25, 93.23±10.4 and 94.41±11.1% for 20 lpm flow rate and 20.78±3.26, 40.39±6.39, 54.85±9.58, 77.28±11.5and 96.53±4.17% for 90 lpm flow rate and for size 0.3-0.5, 0.5-1.0, 1.0-3.0, 3.0-5.0 and 5.0-10.0 µm respectively. Similarly, the mean FE of control green cloth mask (double layer same fabric) is 21.48±1.40, 38.94±4.65, 65.87±8.93, 93.09±5.71 and 98.78±3.50% for 20 lpm flow rate and 19.49±2.98, 30.76±4.03, 61.27±9.73, 81.77±9.16 and 90.23±9.80% for 90 lpm flow rate and for aerosol size 0.3-0.5, 0.5-1.0, 1.0-3.0, 3.0-5.0 and 5.0-10.0 µm respectively.

**Fig. 4.**
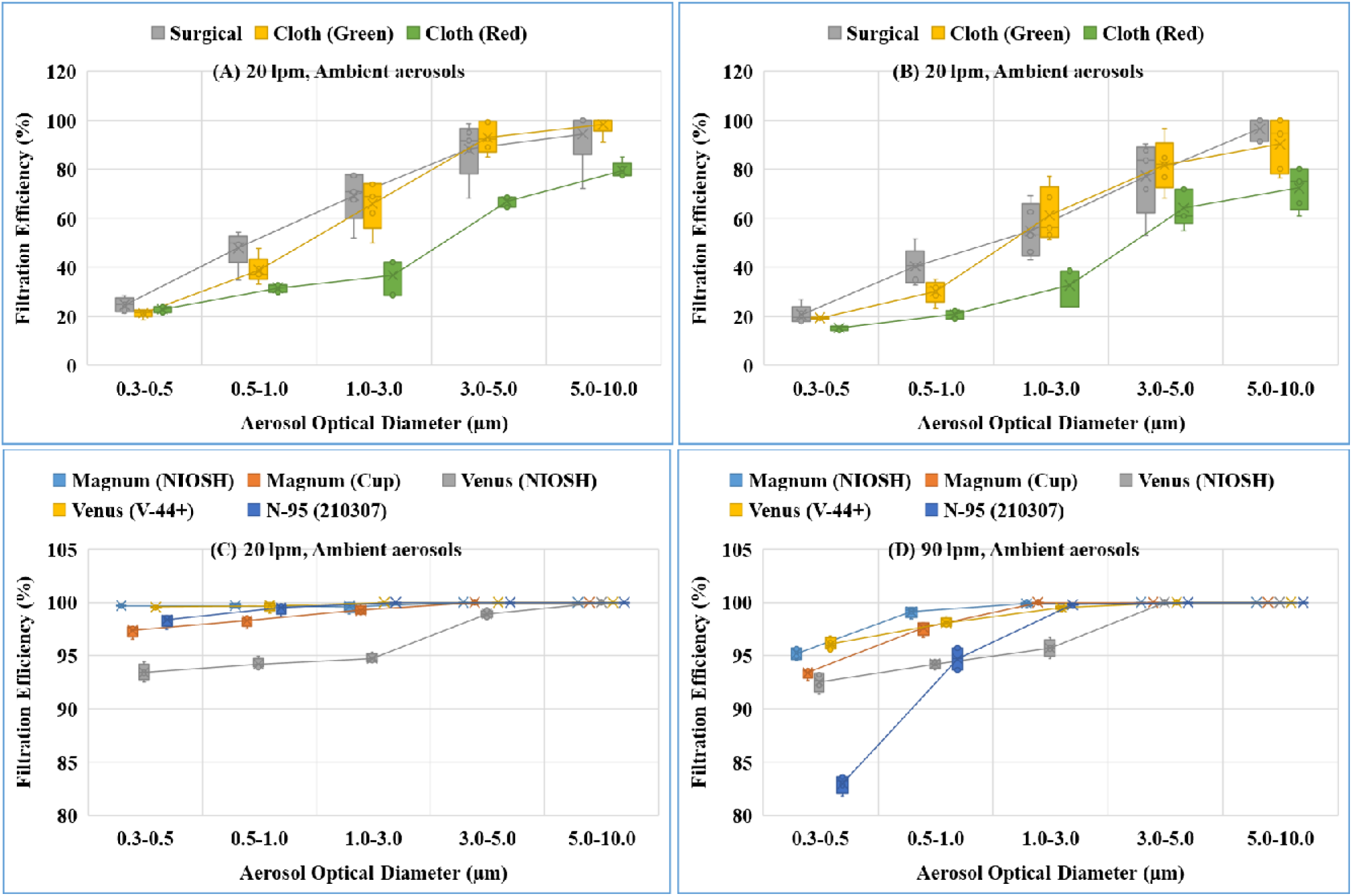
FE of individual control facemasks at a flow rate 20 and 90 lpm for ambient aerosols. (A) and (B) cloth and surgical facemasks (C) and (D) different type certified N-95 facemasks.

The mean FE of control red cloth masks (double layer different fabric) is 21.75±1.12, 31.69±1.35, 36.44±6.45, 66.98±1.90 and 79.96±2.90% for 20 lpm flow rate while for 90 lpm flow rate is 15.38±0.99, 20.86±1.58, 32.76±7.06, 64.01±6.61 and 72.53±7.50% for aerosol size 0.3-0.5, 0.5-1.0, 1.0-3.0, 3.0-5.0 and 5.0-10.0 µm respectively. The FE increases with increase of aerosols optical diameter and data variability increased with the flow rate rising from 20 to 90 lpm and effect is more at larger aerosols optical diameter. The FE of is least for Most Penetrating Particle Size (MPPS: 0.3 - 0.5 µm) for all three type of mask and both flow rates. The FE is more for low flow rate (20 lpm) than high flow rate (90 lpm) and FE of the green cloth masks is more than red cloth mask and less than surgical mask for both flow rates. The red cloth mask is performing slightly lower efficiency than green cloth masks. The higher FE of green cloth mask is attributed to use of same fabric of good thread counts, which helps in trapping the particles efficiently.

As expected, the FE of certified N-95 masks is significantly greater than the surgical and cloth masks for all aerosol measured size ranges. Even MPPS were filtered more than97.37±0.5% except Venus (NIOSH) for all size range of aerosols and for 20 lpm flow rate. Similarly, the FE is larger than 92.54±1.35% except N-95 (210307) face masks for all size range of aerosols andfor 90 lpm flow rate. It can be observed that the FE is more for low flow rate (20 lpm) and less for high flow rate (90 lpm) for all five types of N-95 masks. The certified face masks (N-95) are having electrostatic filters that are efficient at low air flow velocity (flow rate) rather than high airflow velocity (Colbeck and Lazaridis, 2014). The minimum FE is 93.44±0.68% for Venus (NIOSH) and 82.98±1.5% for N-95 (210307) facemask for 20 and 90 lpm respectively. The minimum FE of certified N-95 mask is at MPPS (0.3-0.5 µm) for 20 and 90lpm. The statistical variability in FE is more for cloth masks compared to the surgical mask and it is found to be least for certified N-95 masks. Further, statistical variability in FE of all type of face masks is more for 90 lpm flow rate than 20 lpm.

### 3.4 Evaluation of FE of certified facemasks (N-95) after different sterilization

The N-95 (Magnum NIOSH) face masks are sterilized with the help of dry heat (hot air oven)for 30 and 60 minutes, gamma irradiation for 15 and 25 kGy dose and 10% H_2_O_2_ concentrated liquid soaking after that FE evaluated for two flow rates and shown in Fig.5. It is observed from the Fig.5 that, the FE of N-95 remain same (or slightly decreased) as from control mask within statistical uncertainty after dry heat and H_2_O_2_ sterilization for both flow rates. However, even with slightly reduced FE, the Magnum NIOSH facemasks offered 98.53±0.79% or higher at 20 lpm. At the higher flow rate (90 lpm), the FE is found to be more than 95.16±0.69% for the tested aerosol optical diameter. Further, the FE of Magnum NIOSH facemasks decrease significantly for less than 1.0 µm optical diameter after gamma sterilization (15 and 25 kGy) for both flow conditions. The reduction in FE is more for higher flow rate/face velocity. The similar observation also made by other researchers (Amit et al., 2020 and Cramer et al., 2020). The FE is more for 20 lpm than 90 lpm in all the sterilization conditions. It is known that, the aerosol filtration takes place by five basic mechanisms viz. gravitational settling, inertial impaction, interception, diffusion and electrostatic interaction (Hinds, 1999 and Vincent, 2007). The N95 mask consists of electrostatic filtration media that are capable of capturing and retaining fine aerosol through electrostatic interaction in addition to mechanical processes (Myers and Arnold, 2003). It is known that, least efficiency is associated with particles in the range of 0.3 □ 0.5 µm that is bigger for diffusion and smaller for interception; hence, the 99% efficiency is achieved for this range by electrostatic interaction. When the media loses its charges, the particles are captured only by mechanical processes where the efficiency is reduced from 99±0.09% to 63.73±1.09%. In the case of greater than 1.0 µm particles, the efficiency is not found reduced even after gamma irradiation.

**Fig. 5.**
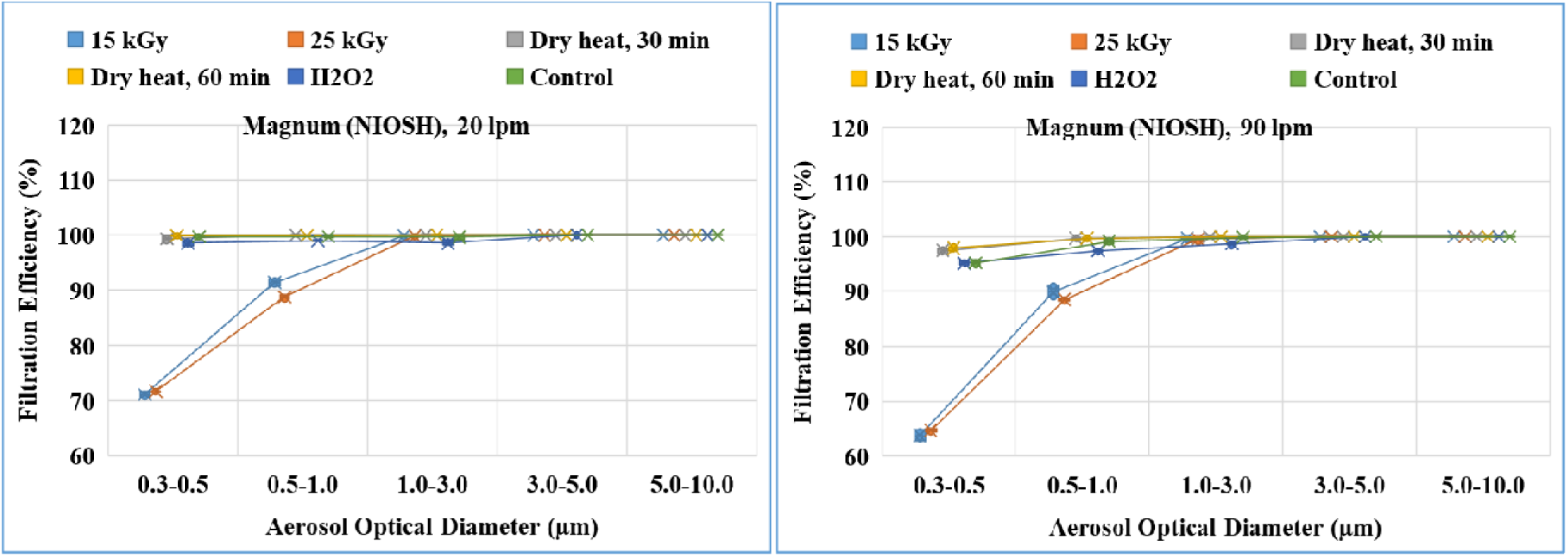
FE of N-95 (Magnum NIOSH) face masks after gamma, dry heat and H_2_O_2_ sterilization for 20 and 90 lpm flow rates.

The *FE* of mechanical filters has least efficiency at MPPS and increases with increase or decrease of aerosol size from MPPS. The FE remain unchanged or more than 95% benchmark for aerosols optical diameter greater than 1.0 µm for tested decontamination methods (dry heat for 30 and 60 min, gamma irradiation for 15 and 25 kGy dose and H_2_O_2_ sterilization).

The fiber structure of magnum N-95 facemaks are examined in the optical microscope (Make and Model: Axioplan 2 Imaging, Metasystems, Germany) under 100X magnification. The image taken by optical microscope for N-95 is shown in Suplemetry Information (SI-1). It is observed that, no observable and significant change in morphology of filter fibre is found with gamma sterilization upto 25 kGy dose. Similar observations were reported by Scanning Electron Microscopy (SEM) up to 61 kGy dose (IAEA technical report, 2020). The International Atomic Energy Agency (IAEA) has recently indicated the same observation that, there is no significant change in the texture of the mask with respect to fit factor of the mask at 24 kGy radiation dose, which is needed to kill viruses and bacteria (http://www-naweb.iaea.org/napc/iachem/working_materials/Technical%20Report%20(Mask%20Reprocessing.pdf).

Further, the N-95 Venus (NIOSH) face masks are sterilized with moisturized heat (Autoclave) for 30 and 60 minutes and 10 % H_2_O_2_ concentrated liquid soaking, after that FE are evaluated for two flow rates and shown in Fig. 6. The FE of Venus (NIOSH) masks increased from 93.44±1.49 to 94.61±1.04 and 92.58±1.59 to 94.70±1.12 for 20 and 90 lpm flow rate respectively after 30 minutes autoclave sterilization. The increase in efficiency is not significant. However, for 60 minute autoclave sterilization, FE decreases from 93.44±1.49 to 92.49±1.89% for 20 lpm flow rate (not significant) and from 92.58±1.59 to 86.50±1.99%for 90 lpm flow rate for optical diameter of aerosols 0.3-0.5µm. In the case of H_2_O_2_ sterilization, FE is found to be increased from 93.44±1.49 to 97.28±0.89% for 20 lpm flow rate and 92.58±1.59 to 97.68±0.81 for 90 lpm flow rate for optical diameter of aerosols 0.3-0.5µm. It is noted that FE is found to be more at 20 lpm than(90 lpm for optical diameter 0.3–0.5, 0.5–1.0 and 1.0–3.0µm.

**Fig. 6.**
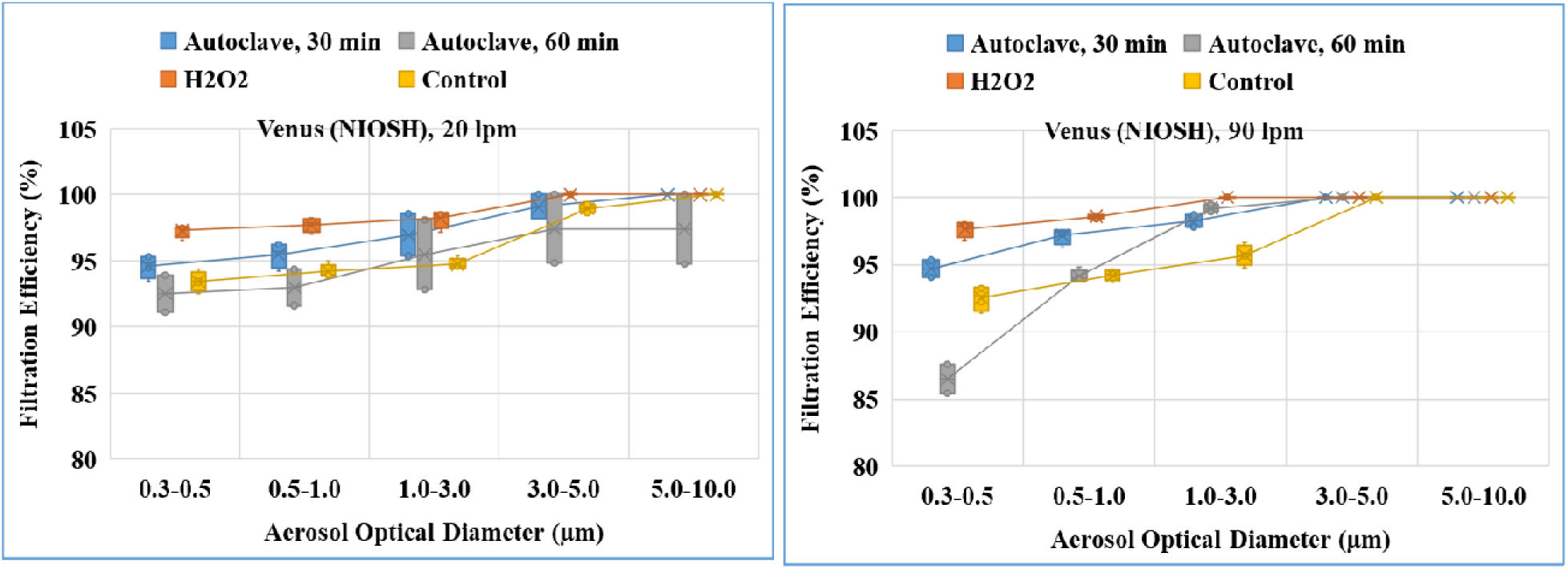
FE of N-95 (Venus NIOSH) face masks after autoclave and H_2_O_2_ sterilization for 20 and 90 lpm flow rates.

The other three types of N-95 face masks (magnum cup shape, V-44+ and 210307) were sterilized with autoclave for 30 minutes only and FE is given in Fig.7. It is observed from the Fig.7 that, the FE of N-95 decreased from control masks for both flow rates. The reduction in FE of N-95 magnum cup shape, V-44+ and 210307 facemasks is from 97.37±0.69 to 96.24±0.52%, 99.56±0.09 to 92.53±0.69% and 98.38±0.87 to 85.27±1.29% respectively for 20 lpm flow rate. Similarly, for 90 lpm flow rate, reduction in FE ofN-95 magnum cup shape, V-44+ and 210307 facemasks is from 93.39±1.69 to 88.20±1.39%, 96.08±0.78 to 89.34±1.02% and 82.98±1.5 to 79.14±2.01% respectively. The reduction of FE after autoclave sterilization is more for magnum cup shape and V-44+ facemasks in high flow conditions compared to the low flow rate condition.

**Fig. 7.**
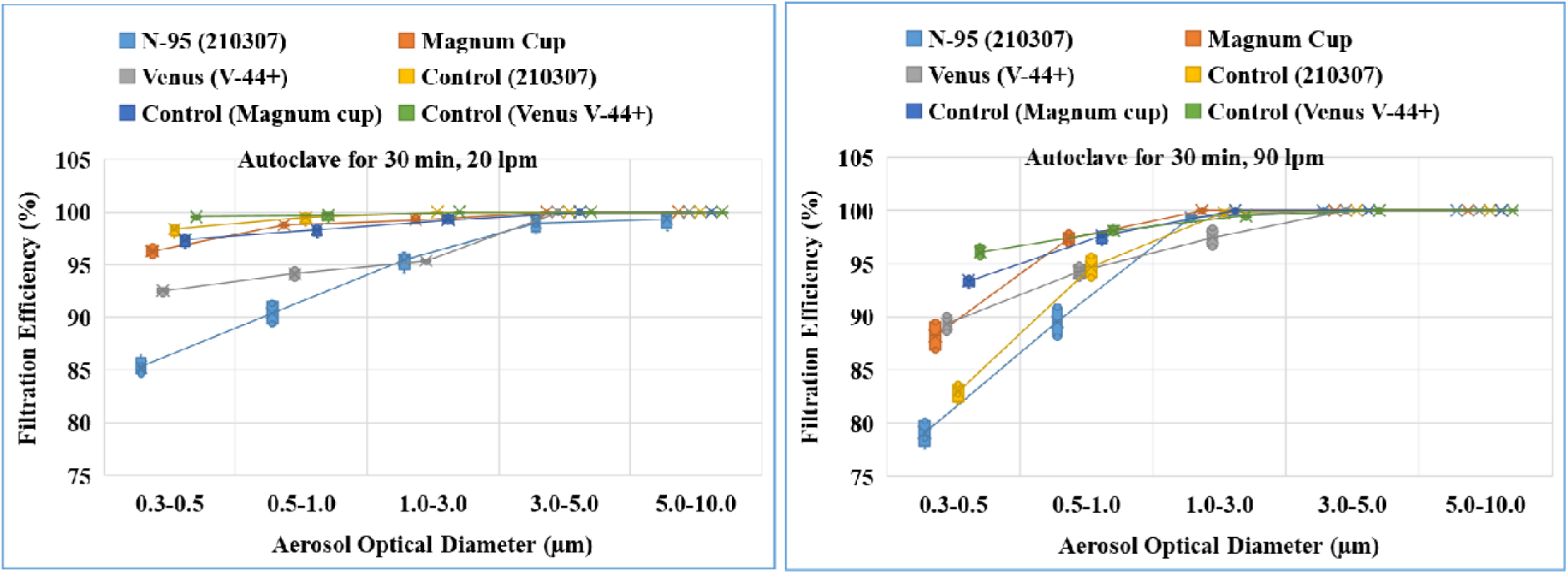
FE of various certified N-95 facemasks after autoclave sterilization for 20 and 90 lpm flow rates.

### 3.6 Evaluation of FE of non-woven fabric masks (surgical equivalent) after different sterilization

The surgical facemasks are sterilized with the help of steam (autoclave) for 30 and 60 minutes, gamma irradiation for 15 and 25kGy dose, 10% H_2_O_2_ concentrated liquid soaking, hot water soaking with and without detergent; after that FE is calculated for two flow rates and shown in Fig.8. As expected from filtration principles, the minimum FE is found in between 0.3–0.5µm for all untreated and processed surgical masks and increases with increase of aerosol optical diameter. The mean FE of untreated surgical masks is 24.72±2.54% and 20.78±3.26% for low and high face velocity respectively. The average *FE* is found to be more or less same after gamma, H_2_O_2_ and hot water sterilization and found to vary from 21.66±2.33 □29.86±1.49% and 16.52±2.44 □22.29±1.59% for 20 lpm and 90 lpm flow rates respectively for ambient aerosols size 0.3 □5.0µm. The average *FE* after autoclave sterilization increases from 24.72±2.54 to 30.34±1.025 for low face velocity while for high face velocity decreases from 20.78±3.26 to 16.28±3.52%. It is noted that, the FE is increases more for 60 minute autoclave sterilization for both flow rate when compared to the 30 minute autoclave.

**Fig. 8.**
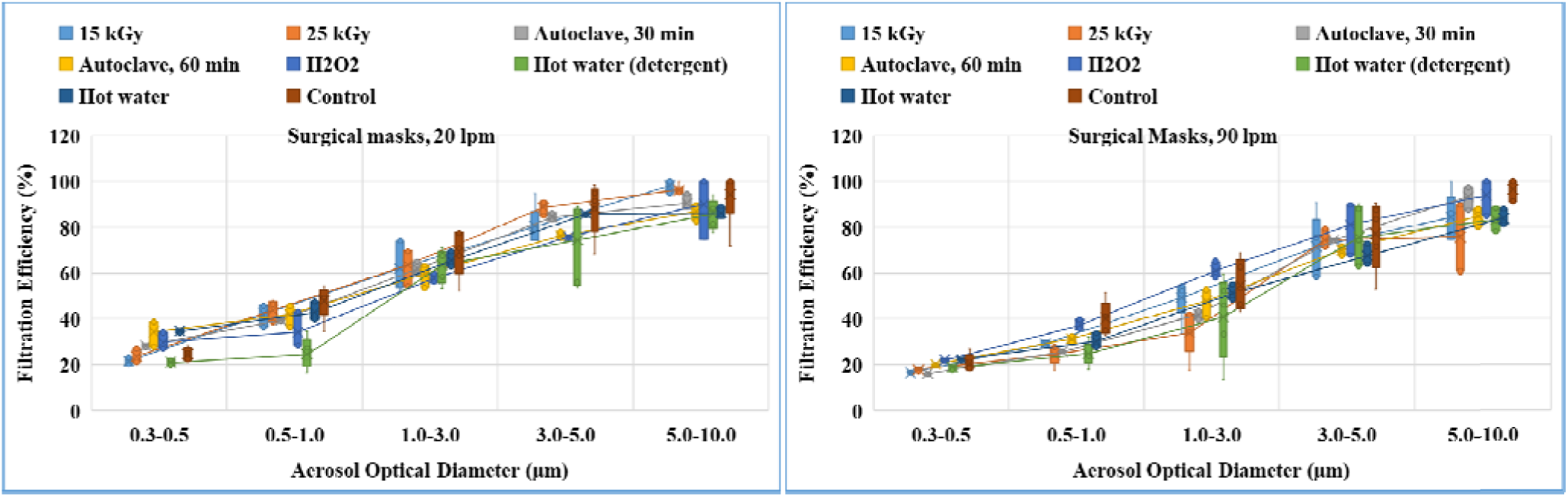
FE of surgical equivalent face masks after gamma, autoclave, H_2_O_2_ sterilization and hot water with and without detergent sterilization for 20 and 90 lpm flow rates.

In the case of hot water with detergent sterilization, the FE reduced slightly from 24.72±2.54 to 21.01±2.62 and 20.78±3.26 to 18.79±2.98% for 20 lpm and 90 lpm flow rate. In general the FE of surgical face mask is around 20-25% for MPPS The spread in FE is low at MPPS and increases with larger aerosols size for both face velocity. The lower efficiency is attributed to density of fibers in the non-woven type masks. However, these masks are effective for the particles in the size range of 3.0 µm and above (about 60% efficiency). The surgical mask consists of three layer viz. repellant the outer colored layer, the filter medium in the middle and absorbent at the innermost layer. The microscopic image of filter media of the control and processed masks (15 kGy and 25 kGy) is given in Supporting Information (SI). It is observed that, the structure of the filter fibre media is bonded togeather by using chemical adhesive, appeared entangled structure and there is no observable change in the structure of the filter fibre after gamma sterilisation.

### 3.7 Evaluation of FE of homemade cloth masks (green and red color) after different sterilization

The homemade double layer cloth facemasks (green color and same fabric)are sterilized with the help of autoclave for 30 and 60 minutes, gamma irradiation for 15 and 25 kGy dose, hot water soaking with and without detergent; after that FE is calculated for two flow rates and shown in Fig.9.

**Fig. 9.**
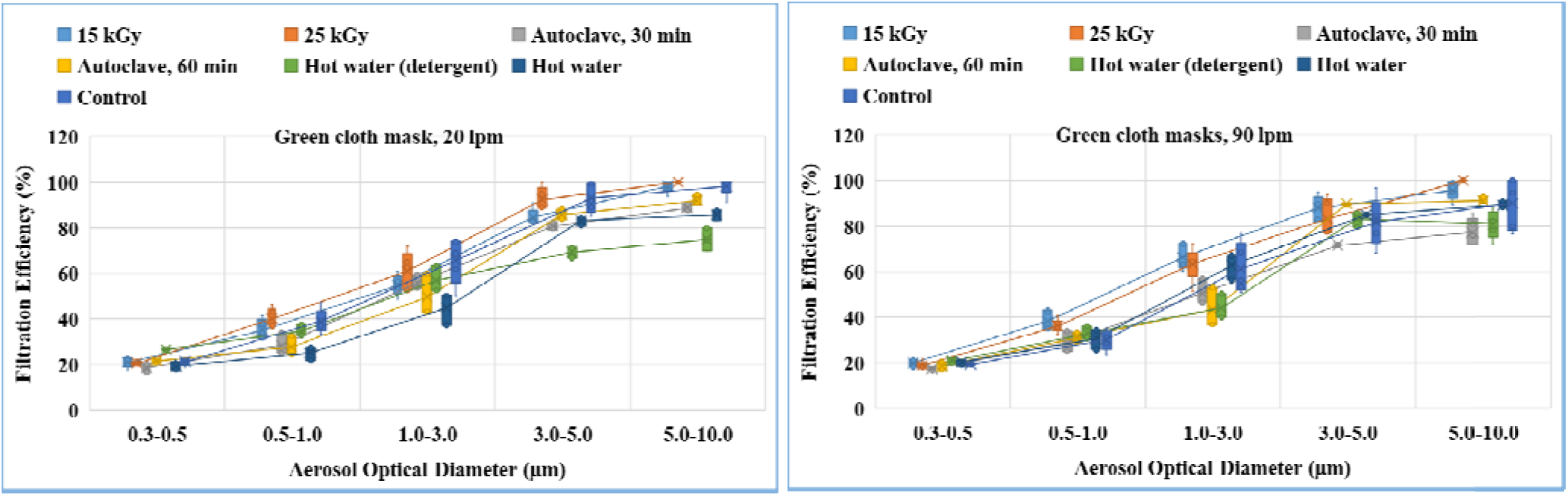
FE of double layer (green cloth same fabric) face masks after gamma, autoclave, and hot water with and without detergent sterilization for 20 and 90 lpm flow rates.

The average *FE* of green cloth masks is found to be more or less same after gamma, autoclave, and hot water (with and without detergent) sterilization and found to vary from 19.69±1.44 □25.86±2.49 % and 17.52±2.84 □20.29±3.29% for 20 lpm and 90 lpm flow rates respectively for ambient aerosols size 0.3□5.0 µm. The average *FE* increases with increase of aerosols optical diameter for both face velocity before and after sterilization condition. The homemade double layer cloth facemasks (red color and different fabric)are sterilized with the help of autoclave for 30 and 60 minutes and hot water (with and without detergent); after that FE are calculated for two flow rates and shown in Fig.10.

**Fig. 10.**
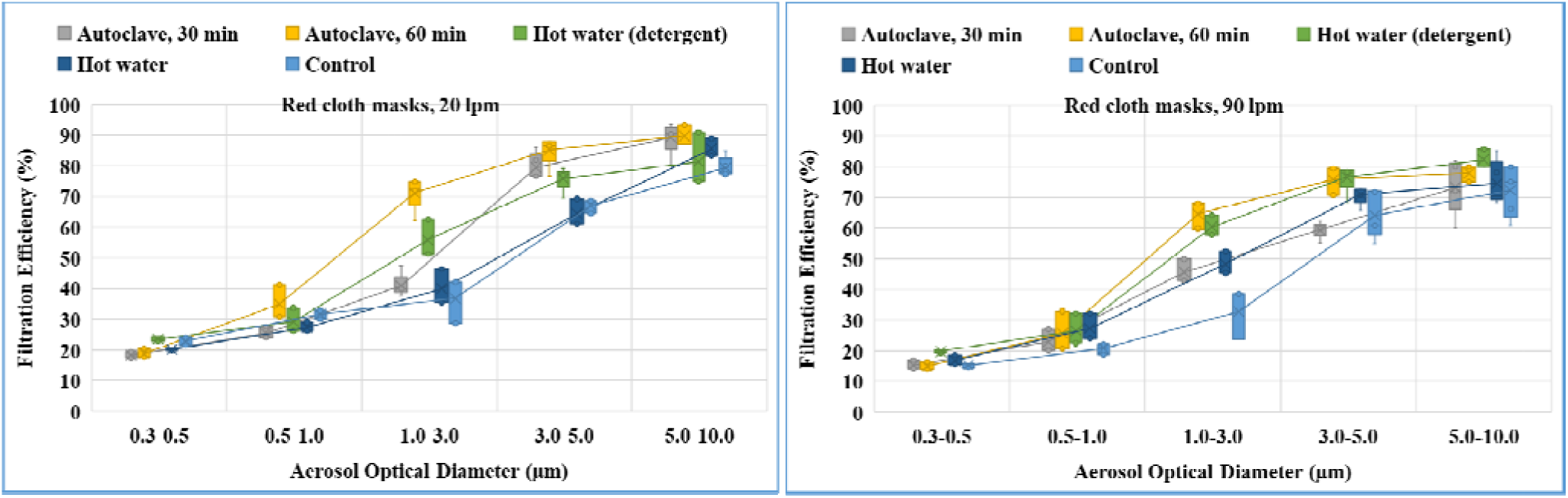
FE of double layer (red cloth different fabric) face masks after autoclave, hot water with and without detergent sterilization for 20 and 90 lpm flow rates.

The average *FE* of red cloth masks is found to be more or less same (18.28±1.56 – 23.52±2.79%) for 20 lpm and (14.98±1.87 – 19.83±2.09%) aerosol size 0.3-0.5 µm. Similar to green colored same fabric mask, The mean FE increase as aerosol optical diameter increases from 0.5-1.0 to 5.0 −10.0 µm for both sterilized and non-sterilized conditions. It is noted that, the FE is relative less for red colored different layered cloth masks than with that of green colored same fabric double layered mask. The reason is attributed to texture (less thread counts) of second layer in the red colored mask. The uncertainty in *FE* is significantly more for cloth and surgical masks in case greater than 0.3 – 0.5 µm aerosols compared to certified N95 masks probably indicating the texture/material quality of masks in the manufacturing stage.

### 3.8 Comparison of FE of various type of masks after different sterilization for ambient and PAO aerosols

The performance of various types of masks after different sterilization methods has been evaluated by laboratory generated aerosols (PAO) and FE has been compared from ambient aerosols. This type of analysis may be very useful for some specific context where a person exposed to the higher aerosols concentration. For example, the average number of aerosols generated per cough by Influenza patient is 7.5*10^4^ and count median diameter (CMD) of cough generated aerosols/droplet were in between 0.6 to 0.9 µm with Geometric Standard Deviation (GSD) 1.53 to 2.28 (Lindsley *et al*., 2012). Similarly, another recent study suggests that, the average number of droplet/aerosols expelled per cough by a person having respiratory infection is 4.9*10^6^ with most of the aerosols are in sub micron (Lee *et al*., 2019). Another study shows that 80% of droplets/aerosols are centered in the range of 0.74 □ 2.12 µm during coughing and sneezing (Yang *et al*., 2007). In this context, the filtering efficiency of the mask needs to be tested for high aerosols concentration (10^5^ □ 10^6^ #/L).Table 5 shows the FE of various types of facemasks for ambient and PAO aerosols after different sterilization methods, for the two flow rate conditions and for MPPS (0.3-0.5) aerosol optical diameter. It is observed from the Table 5 that, certified N-95 mask shows slightly greater FE for laboratory generated aerosols (high aerosols concentration) than ambient aerosols even after sterilization for both flow rate conditions.

**Table 5.**
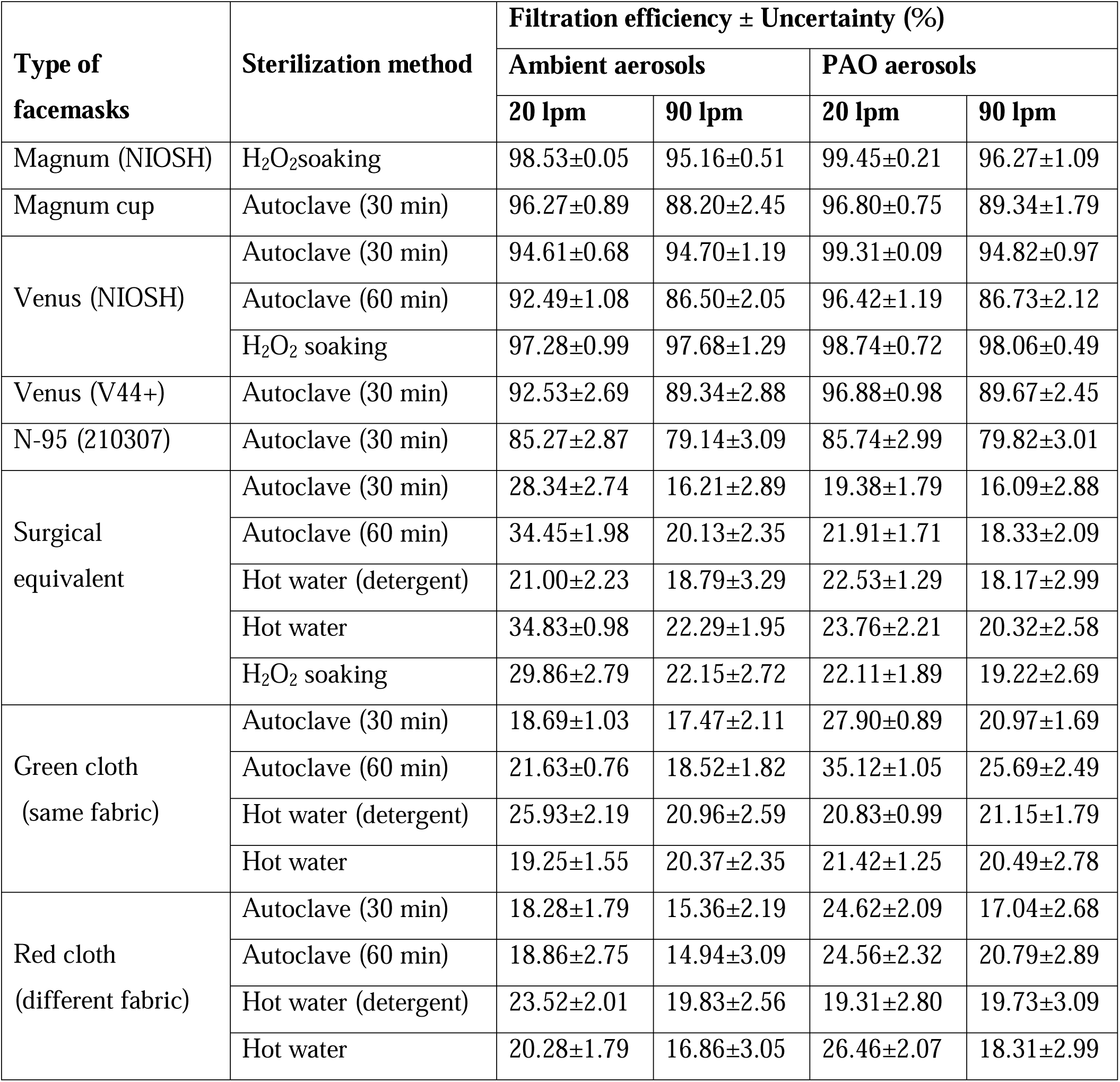
Comparison of FE of various types of facemasks for ambient and PAO aerosols after different sterilization.

| Type of facemasks | Sterilization method | Filtration efficiency ± Uncertainty (%) | | | |
| --- | --- | --- | --- | --- | --- |
|  |  | Ambient aerosols |  | PAO aerosols |  |
|  |  | 20 lpm | 90 lpm | 20 lpm | 90 lpm |
| Magnum (NIOSH) | H<sub>2</sub>O<sub>2</sub>soaking | 98.53±0.05 | 95.16±0.51 | 99.45±0.21 | 96.27±1.09 |
| Magnum cup | Autoclave (30 min) | 96.27±0.89 | 88.20±2.45 | 96.80±0.75 | 89.34±1.79 |
| Venus (NIOSH) | Autoclave (30 min) | 94.61±0.68 | 94.70±1.19 | 99.31±0.09 | 94.82±0.97 |
| | Autoclave (60 min) | 92.49±1.08 | 86.50±2.05 | 96.42±1.19 | 86.73±2.12 |
| | H<sub>2</sub>O<sub>2</sub> soaking | 97.28±0.99 | 97.68±1.29 | 98.74±0.72 | 98.06±0.49 |
| Venus (V44+) | Autoclave (30 min) | 92.53±2.69 | 89.34±2.88 | 96.88±0.98 | 89.67±2.45 |
| N-95 (210307) | Autoclave (30 min) | 85.27±2.87 | 79.14±3.09 | 85.74±2.99 | 79.82±3.01 |
| Surgical equivalent | Autoclave (30 min) | 28.34±2.74 | 16.21±2.89 | 19.38±1.79 | 16.09±2.88 |
| | Autoclave (60 min) | 34.45±1.98 | 20.13±2.35 | 21.91±1.71 | 18.33±2.09 |
| | Hot water (detergent) | 21.00±2.23 | 18.79±3.29 | 22.53±1.29 | 18.17±2.99 |
| | Hot water | 34.83±0.98 | 22.29±1.95 | 23.76±2.21 | 20.32±2.58 |
| | H<sub>2</sub>O<sub>2</sub> soaking | 29.86±2.79 | 22.15±2.72 | 22.11±1.89 | 19.22±2.69 |
| Green cloth (same fabric) | Autoclave (30 min) | 18.69±1.03 | 17.47±2.11 | 27.90±0.89 | 20.97±1.69 |
| | Autoclave (60 min) | 21.63±0.76 | 18.52±1.82 | 35.12±1.05 | 25.69±2.49 |
| | Hot water (detergent) | 25.93±2.19 | 20.96±2.59 | 20.83±0.99 | 21.15±1.79 |
| | Hot water | 19.25±1.55 | 20.37±2.35 | 21.42±1.25 | 20.49±2.78 |
| Red cloth (different fabric) | Autoclave (30 min) | 18.28±1.79 | 15.36±2.19 | 24.62±2.09 | 17.04±2.68 |
| | Autoclave (60 min) | 18.86±2.75 | 14.94±3.09 | 24.56±2.32 | 20.79±2.89 |
| | Hot water (detergent) | 23.52±2.01 | 19.83±2.56 | 19.31±2.80 | 19.73±3.09 |
| | Hot water | 20.28±1.79 | 16.86±3.05 | 26.46±2.07 | 18.31±2.99 |

The FE of surgical mask is found to be reduced for high aerosols concentration. However, the cloth masks (green and red color), the FE has been increased for laboratory generated aerosols. It is also to be noted that the FE of all three types of mask is less for 90 lpm flow rate when compared to the 20lpm.This is due to higher filtration capability for sub-micron sized aerosols by diffusion and interception at low flow rate.

## 5.0 Summary and conclusion

Three types of facemasks viz. certified N-95, surgical equivalent and self-made double layer cloth masks have been tested for particulate *FE* and pressure drop across the masks for two-flow rate condition viz. 20 and 90 lpm before and after various sterilization using ambient aerosols in the size range of 0.3 □ 10 µm and laboratory generated aerosols (PAO) of size 0.3 −0.5 µm. As expected, the pressure drop increases with increasing face velocity, this reflect that, it is harder to breathe through facemask when respiratory flow is large. The measured pressure drops during masks testing were found to be accepted range (the inhalation and exhalation resistance limit viz. 35 and 25 mm of H_2_O respectively). The *FE* of surgical masks is more or less same as green colored cloth masks (double layer of same fabric) for 0.3 to 10 µm optical diameters for 20 and 90 lpm flow rate. However, the *FE* of red cloth masks (double layer with different fabric) is less compared to the surgical masks. The *FE* of cloth and surgical masks increases with increase of optical diameter from MPPS and the efficiency is less at high flow rate compared to the low flow rate. In the case of N-95 mask, the FE is found to be more than 95% for control masks, for both flow rates and ambient aerosols (0.3□10µm). The FE reduces to about 70% for most penetrating aerosols (0.3 µm) after gamma sterilization for 20 lpm flow rate and still lesser with higher flow rate (90 lpm). The results found that the cloth masks that underwent autoclave for duration of 60 minutes have shown better efficiency. The increased in *FE* of surgical masks for autoclave of 60 minutes duration is because the mask is subjected to steam for prolonged time, which shrink the fiber of the facemask but same time destroying of the microbes and melted the elastic cords of surgical masks to a slight extent.

Face masks can provide additional protection when some measures, particularly social distancing is difficult to maintain. The effectiveness of facemasks depends on consistent and proper use. Though the surgical masks have slightly better efficiency than the double layer cloth masks, we assume that the efficiency can be matched with it if the cloth masks have preferably three layers preferably with fabric having good thread counts (fabric with adequate droplet blocking efficiency). Thus, we can recommend the use of cloth mask for the common public provided there are automatic hand sanitizers installed before entering a crowded place or an office. It has to be made sure that the cloth masks are sterilized with any of the method above (preferably hot water sterilization in houses) before subsequent uses. The *FE* of N-95 masks is slightly decreases after autoclave for 30 and 60 minutes duration because the mask is subjected to steam of 120°C for prolonged time which results in destroying of the microbes. At the same time, the process melted the elastic cords of the N-95 and surgical masks to a slight extent. The *FE* of the N-95 masks remain almost the same compared to the control condition after the H_2_O_2_ sterilization but faded the names in ink in the masks. The FE of certified masks more or less same before and after dry heat sterilization for 30 and 60 minutes duration. Thus, it is recommended that, the N-95 masks can be sterilized in a few times with dry heat and H_2_O_2_ vapors. Further, the autoclave sterilization methods can be also used but temperature of stream should be kept less (< 100°C) so that the elastic cords of the masks remained in good condition.

## Recommendations

We highlight few recommendations from the present studies:

- The breathability test (pressure drop) and filtering efficiency conveys that, the cloth masks with double or triple layer with tight weaves, low porosity and high thread count are potential substitute for medical or surgical masks (not for certified N-95 masks).
- The cotton mask is a potential substitute for the public instead of a surgical mask due to its cost effectiveness and it could be reused even after decontamination by any above methods (preferably 80-90°C hot water wash).
- We recommend that hot water washing with or without detergent is quite good for sterilization of cloth masks which is very much affordable for the common public and it is very effective.
- N-95 masks, which are made of electrets filtering media, are not recommended for sterilisation or decontamination by ionising radiation, it will compromise the filtering efficiency. However, it is recommended that, masks can be sterilized in a few times with dry heat and H_2_O_2_ vapors.

## Data Availability

Yes

## 6.0 Acknowledgement

The authors acknowledge the Dr. S. Athamalingam, Associate Director, Health, Safety and Environmental Group (HSEG), Dr. R. Venkatesan, Head, Radiological and Environmental Safety Division (RESD) and Dr. C. V. Srinivas, Head, Radiological Impact Assessment Section (RIAS) for their encouragement and support to carry out this work.

