## Supplemental Information for "Evaluation of filtration efficacy of various types of facemasks using ambient and PAO aerosols following with different sterilization methods"

The dimension of each image is 65 µm x 65 µm and the filter fiber diameter is in sub-micrometer range for N-95 mask while micrometer ranges for surgical mask.

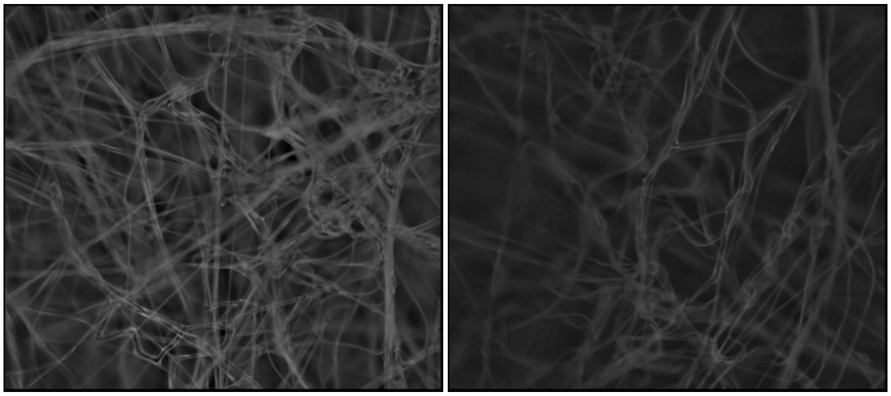

(A) Control

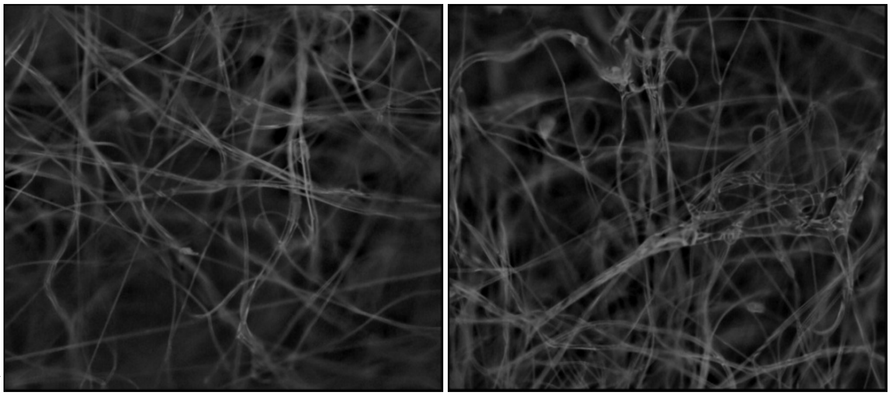

(B) 15 kGy

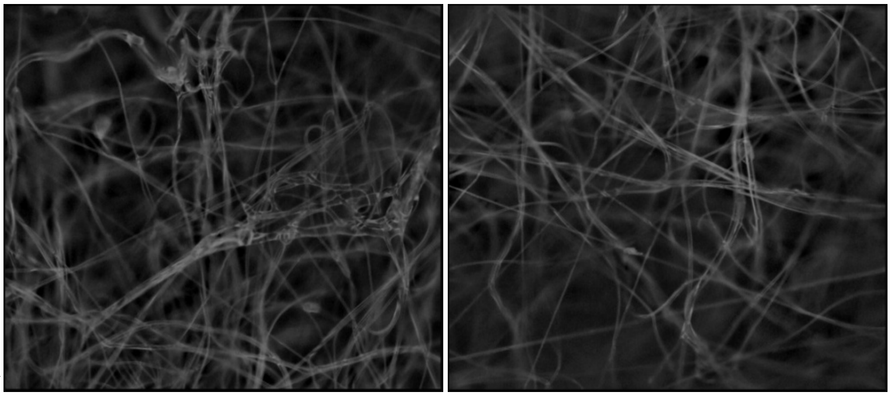

(C) 25 kGy

**Fig.1. Optical microscope image of Filter layers of magnum N-95 mask (a) control, (b) 15 kGy and (c) 25 kGy.**

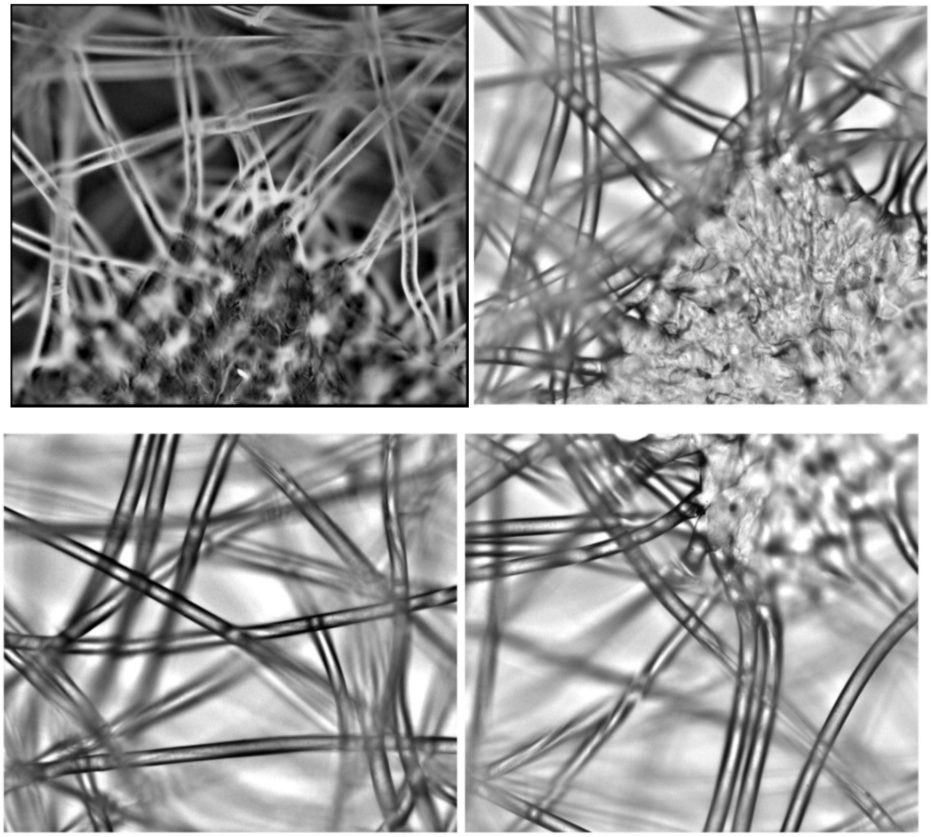

**(A) Control**

**(B) 15 kGy**

**(C) 25 kGy** 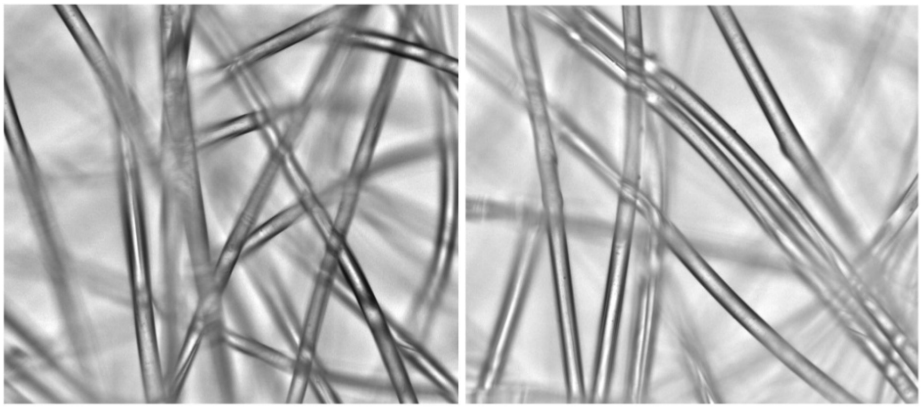

**Fig.2 Optical microscope image of filter layers of surgical mask (a) control, (b) 15 kGy and (c) 25 kGy.**
